# An Objective Search for Unrecognized Bias in Validated COVID-19 Prediction Models

**DOI:** 10.1101/2021.10.28.21265629

**Authors:** Hossein Estiri, Zachary H Strasser, Sina Rashidian, Jeffery G Klann, Kavishwar B Wagholikar, Thomas H McCoy, Shawn N Murphy

## Abstract

The growing recognition of algorithmic bias has spurred discussions about fairness in artificial intelligence (AI) / machine learning (ML) algorithms. The increasing translation of predictive models into clinical practice brings an increased risk of direct harm from algorithmic bias; however, bias remains incompletely measured in many medical AI applications. Using data from over 56 thousand Mass General Brigham (MGB) patients with confirmed severe acute respiratory syndrome coronavirus 2 (SARS-CoV-2), we evaluate unrecognized bias in four AI models developed during the early months of the pandemic in Boston, Massachusetts that predict risks of hospital admission, ICU admission, mechanical ventilation, and death after a SARS-CoV-2 infection purely based on their pre-infection longitudinal medical records.

We discuss that while a model can be biased against certain protected groups (i.e., perform worse) in certain tasks, it can be at the same time biased towards another protected group (i.e., perform better). As such, current bias evaluation studies may lack a full depiction of the variable effects of a model on its subpopulations.

If the goal is to make a change in a positive way, the underlying roots of bias need to be fully explored in medical AI. Only a holistic evaluation, a diligent search for unrecognized bias, can provide enough information for an unbiased judgment of AI bias that can invigorate follow-up investigations on identifying the underlying roots of bias and ultimately make a change.

## Introduction

The healthcare research and industry have been increasingly progressive on the translation and implementation of artificial intelligence (AI)/machine learning (ML) to improve outcomes and lower costs. Diligently identifying and addressing biases in AI/ML algorithms (hereafter, referred to as “algorithms”) have garnered widespread public attention as pressing ethical and technical challenges.^1–5^ For instance, there is growing concern that algorithms may import and/or exacerbate ethno-racial and gender disparities/inequities through the data used to train them, due to their math, or the people who develop them.^6,7^

The costs of deploying algorithms in healthcare carelessly could exacerbate the very health inequalities society is working to address.^8,9^ The Algorithmic Accountability Act of 2019^10^ requires businesses to evaluate risks associated with algorithm fairness and bias.^11^ Nevertheless, regulating algorithm biases in healthcare remains a difficult task. Eminent cases of algorithm bias have been documented, for example, in facial recognition and natural language processing (NLP) algorithms. Facial recognition systems, for instance, that are being increasingly utilized in law enforcement often perform poorly in recognizing faces of women and Black individuals.^12–14^ In NLP, language is often encoded in gendered formats.^15–17^

These issues are relevant in the healthcare domain, with different caveats at the bench and at the bedside. The demographics (e.g., ethnic, racial) of the patients used to train algorithms is often unknown for external evaluation.^8^ As a result, algorithms have been observed to produce inferior performance in detecting melanoma and health risk estimation in disadvantaged poorer African-American populations.^7,18,19^ Such biases in healthcare may be caused by missing data (e.g., higher rates of missingness in minority populations due to decreased access to healthcare or lower healthcare utilization), observational error, misapplication, and overfitting due to small sample sizes or limited population and practice heterogeneity. ^5,20–22^

In general, algorithm biases can be categorized under statistical and social. Statistical bias, which is common in predictive algorithms, refers to algorithmic inaccuracies in producing estimates that significantly differ from the underlying truth. Social bias embodies systemic inequities in care delivery leading to suboptimal health outcomes for certain populations. ^23^ Social bias can underly statistical bias. In healthcare, we could have a third category of “latent” biases, which refers to increases in social or statistical biases over time due to the complexities of the healthcare processes.^5^

Despite the eminent work in other fields, bias often remains unmeasured or partially measured in healthcare domains. Most published research articles only provide information about very few performance metrics -- mostly through measures of algorithm’s discrimination power, such as the Area Under the Receiving Operating Characteristics Curve (AUROC). The few studies that officially aim at addressing bias, usually utilize single measures (e.g., model calibration^7^) that do not portray a holistic picture of the story on bias. Proper evaluation of bias in medical AI requires a holistic evaluation with a diligent search for unrecognized bias, which can invigorate follow-up investigations to identify the underlying roots of bias.

The COVID-19 pandemic resulted in the generation of new data and data infrastructures related to both pandemic illness and healthcare more broadly. In this paper, we evaluate unrecognized statistical and latent biases from multiple perspectives using a set of AI prediction models developed and validated retrospectively during the first six months of the COVID-19 pandemic.

These models predict risks of mortality, hospitalization, ICU admission, and ventilation due to COVID-19 infection.^24^ We characterize the evaluation of bias into model-level metrics and propose a new approach for evaluating bias from an individual level. We argue that proper evaluation of bias in medical AI requires a holistic approach that can invigorate follow-up investigations for identifying the underlying roots of bias, rather than providing a partial perspective that may not lead to constructive improvement.

## Methods

We study unrecognized bias in four validated prediction models of COVID-19 outcomes to investigate whether a) the models were biased when developed (we refer to this as a retrospective evaluation) and b) the bias changed over time when applying the models on new COVID-19 patients who were infected after the models were trained (we refer to this as a prospective evaluation).

We recently developed an AI pipeline, MLHO, for predicting risks of hospital admission, ICU admission, invasive ventilation, and death in patients who were infected with COVID-19, only using the data from prior to the COVID-19 infection.^24,25^ MLHO models were developed on data from the first six months of the pandemic in Boston -- i.e., between March and October of 2020. MLHO produces and evaluates several models using different classification algorithms and train-test sampling iterations -- for more details see ^24^. For each outcome, MLHO developed several models using different classification algorithms and/or train-test sampling. To evaluate bias, we first select the top 10 models for each outcome based on their retrospective AUROC -- i.e., the discrimination metric obtained on the test set when the models were tested retrospectively. Then we apply the models to data from the retrospective cohort (who were infected with COVID-19 after the models were trained) to evaluate retrospective bias as a baseline. In addition to retrospective evaluations, we also perform prospective bias evaluations by applying these models to patient data from the subsequent 10 months to evaluate temporal changes in discrimination, accuracy, and reliability metrics (Figure 1). We evaluate bias by race, ethnicity, gender, and across time, by comparing the multiple bias metrics against the overall models, which were trained on all patients.

**Figure 1.**
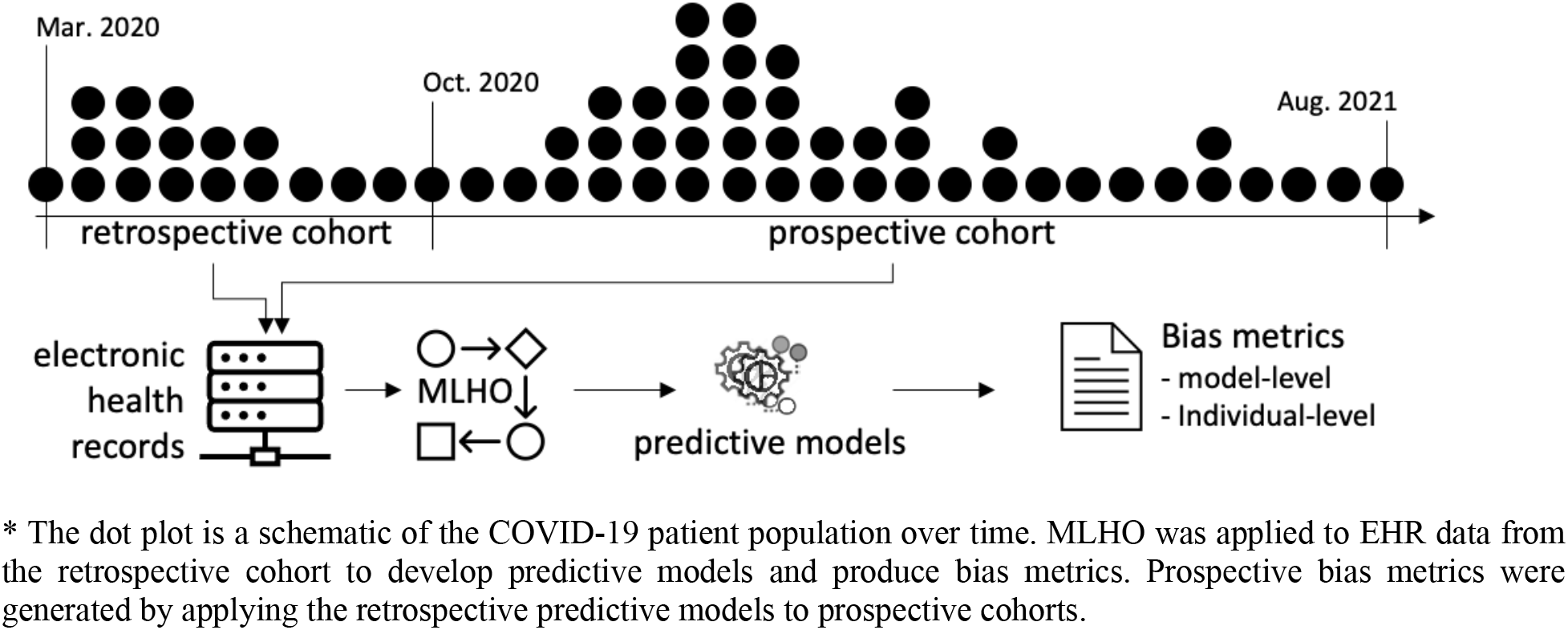
Generating bias metrics from MLHO models using EHR data from retrospective and prospective COVID-19 cohorts.

## Data

Data from 56,590 Mass General Brigham (MGB) patients, with a positive reverse transcription-polymerase chain reaction (RT-PCR) test for SARS-CoV-2 between March 2020 and September 2021 were analyzed (Table 1S). Features utilized in MLHO models included transitive sequential patterns,^26,27^ where we mined sequences of EHR diagnoses, procedures, and medications extracted from these patients’ electronic health records from between 2016 and 14 days before their positive reverse transcription-polymerase chain reaction (RT-PCR) test.

### Measuring bias

To measure bias, we adapt the definitions in ^28–30^ where an unbiased algorithm reflects the same likelihood of the outcome, irrespective of the individual’s group membership, *R*. That is, for any predicted probability score *ŷ*, people in all groups *R* must have equal probability of correctly belonging to the positive class -- for example, *P*(*Y* = 1|*Ŷ* = *ŷ, R* = *black*) = *P*(*Y* = 1|*Ŷ* = *ŷ, R* = *white*).

### MLHO’s performance metrics

MLHO is equipped with functionality to provide a comprehensive evaluation of model performance from different standpoints, including both model-level and individual-level bias.

#### Model-level metrics

The model-level performance metrics in MLHO provide an overall description of the model’s performance, including standard metrics for discrimination, accuracy, and reliability (a.k.a., calibration). For discrimination, in this study, we use the widely used AUROC. Several model-level metrics are also available to evaluate the model’s accuracy such as the Brier score,^31^ which is the mean squared error between the observed outcome and the estimated probabilities for the outcome, including components of both discrimination and calibration.^32^ We break down the AUC and Brier metrics retrospectively in aggregate, and prospectively by month. To compare model-level metrics, we apply the Wilcoxon rank-sum test with Benjamini, Hochberg, and Yekutieli p-value correction.^33^

Reliability is a key factor in AI/ML models’ utility in clinical care, which is also known as calibration. Reliability refers to the extent to which the observed value of an outcome *Y* matches the risk score *R* produced by a predictive model.^7,29^ Several measures have been recommended for measuring model calibration in binary classifiers. For a review of the available techniques, see Huang et al. (2020).^34^ However, many medical AI/ML models developed in healthcare settings ignore reliability and only report discrimination power although the AUROC, also known as the concordance statistic or c-statistic.^35^ MLHO’s performance report provides the ability to assess the models’ reliability for clinical interpretation using diagnostic reliability diagrams. The diagnostic reliability diagrams are produced from the raw predicted probabilities computed by each algorithm (X-axis) against the true probabilities of patients falling under probability bins (Y-axis). In a reliable model, the reliability diagrams appear along the main diagonal—the closer to the line, the more reliable. To evaluate diagnostic reliability diagrams, we compare the retrospective performance with aggregated prospective performance -- i.e., we do not break down this measure by month prospectively.

#### Individual-level metric

In contrast to model-level metrics that provide an overall description of the model’s performance, MLHO also provides the capability for evaluating model performance at an individual level, when the variable of interest is continuous. This is important when assessing whether a model is biased against an individual, for example, an older patient or a sicker patient (i.e. having more medical encounters). To do that, MLHO computes and records the Mean Absolute Error (MAE) for each patient that can be visualized to illustrate changes across continuous indices of interest. MAE is the absolute distance between the computed probability of the outcome to the actual outcome.

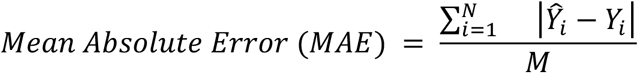

Where *M* is the number of models (10 models here), *Ŷ*_*i*_ is the predicted probability for patient *i* and *Y*_*i*_ is the observed outcome for patient *i*.

To visualize the MAE patterns, we plot the continuous variables (in this study, age) on the X-axis and the MAE on the Y-axis and fit a generalized additive model (GAM) with integrated smoothness^36^ from R package ^37^.

## Results

Data from 56,590 patients with a positive COVID test were analyzed. Over 15,000 of these patients constituted the retrospective cohort -- i.e., whose data was used to train and test the retrospective models. More than 41,000 of the patients were infected between November 2020 and August 2021, who composed our prospective cohort. Figure 1S and Table 1S provide a demographic breakdown of the patient population over time.

### Model-level evaluations

Figures 2 and 3 illustrate temporal changes in the AUROC and Brier scores across the models. The models’ performance metrics remained stable until June 2021. That is, the models that were developed with data from March to September 2020 were still able to perform similarly up until May-June 2021. Starting June 2021, both AUROC and Brier scores exhibit variabilities, in general providing better discrimination power for Hispanic and female COVID-19 patients compared with male patients. In other words, the models did not demonstrate temporal bias until June 2021, when applied prospectively.

**Figure 2.**
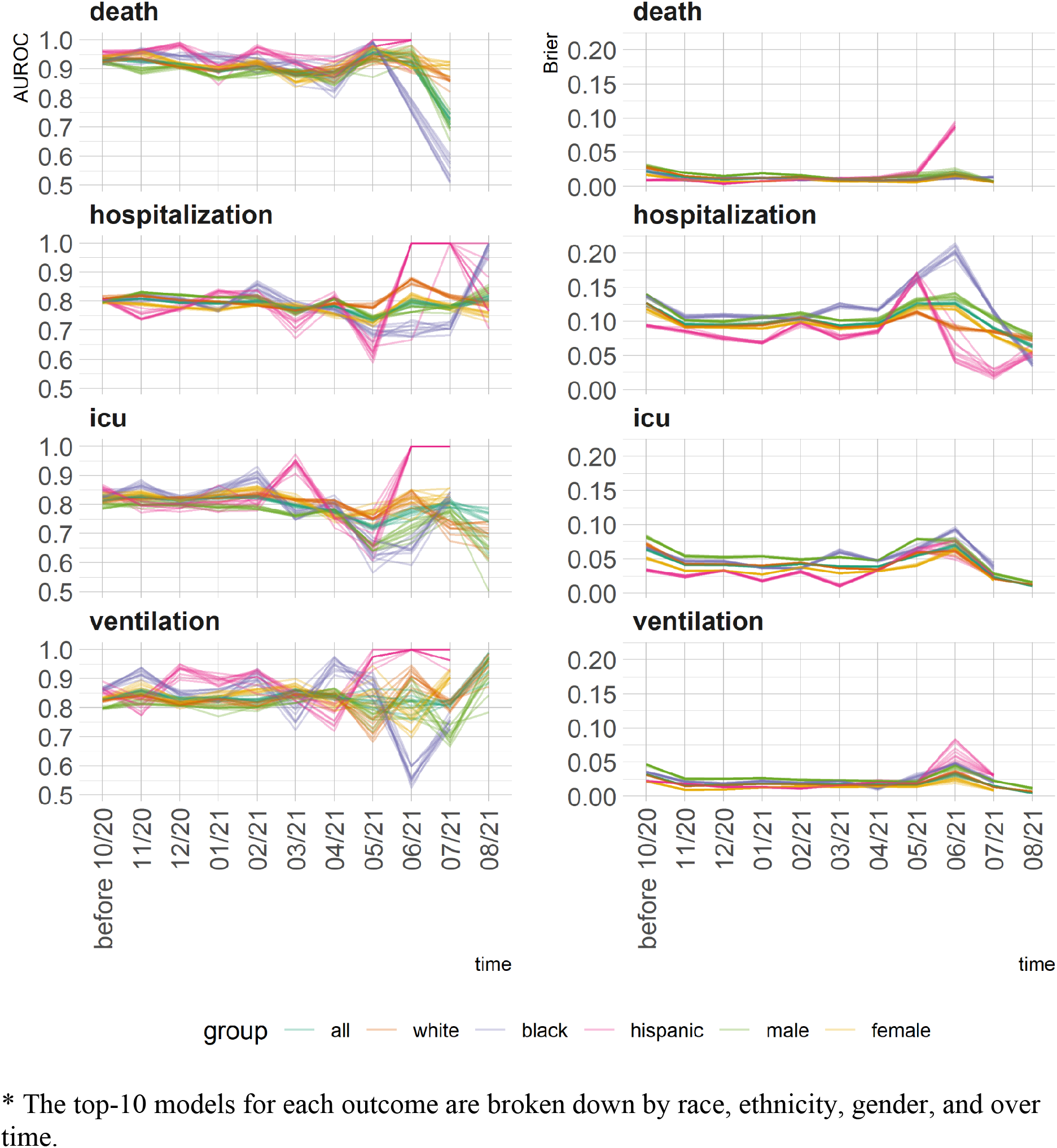
Changes in the 2 model-level metrics for discrimination (AUROC -- left panels) and error (Brier score -- right panel) by group and over time.

**Figure 3.**
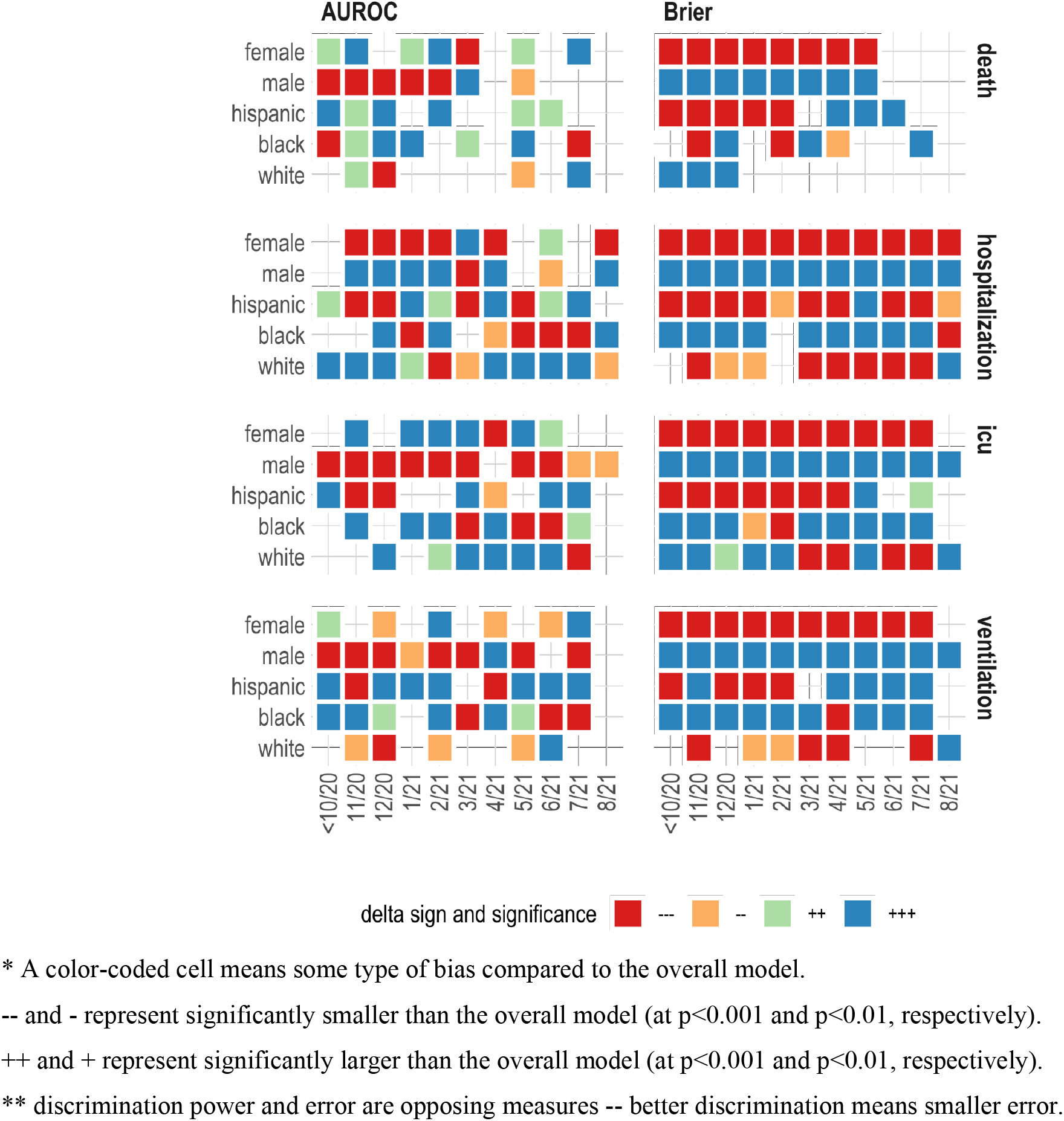
Comparing model-level performance metrics using the Wilcoxon rank-sum test

The models that were developed with data from March to September 2020, provided relatively stable predictive performance prospectively up until May-June 2021. Despite the increased variability, the prospective modeling performance remained high for predicting hospitalization and the need for mechanical ventilators.

To facilitate understanding Figure 2, we provide Figure 3 in which we compare model-level performance metrics using the Wilcoxon rank-sum test. The figure combines an illustration of statistical significance and sign for comparing a given metric for a demographic group to the overall model at a point in time. For example, +++ under AUROC for the female patients in November 2020 means that the AUROC was higher for females compared with the overall model and the difference was statistically significant at p< 0.001.

Figure 4 presents diagnostic reliability diagrams, broken down by demographic group and temporal direction of the evaluation (retrospective vs. prospective). Any divergence from the diagonal line in the diagnostic reliability diagrams means lower reliability. The diagrams show that, retrospectively, models’ predicted probabilities were similar across groups for predicting mortality, hospitalization, and ventilation. Prospectively, between-group variability in models fared similarly, although the uncalibrated predicted probabilities were less reliable for mortality prediction, specifically, among hispanic, black, and female patients.

**Figure 4.**
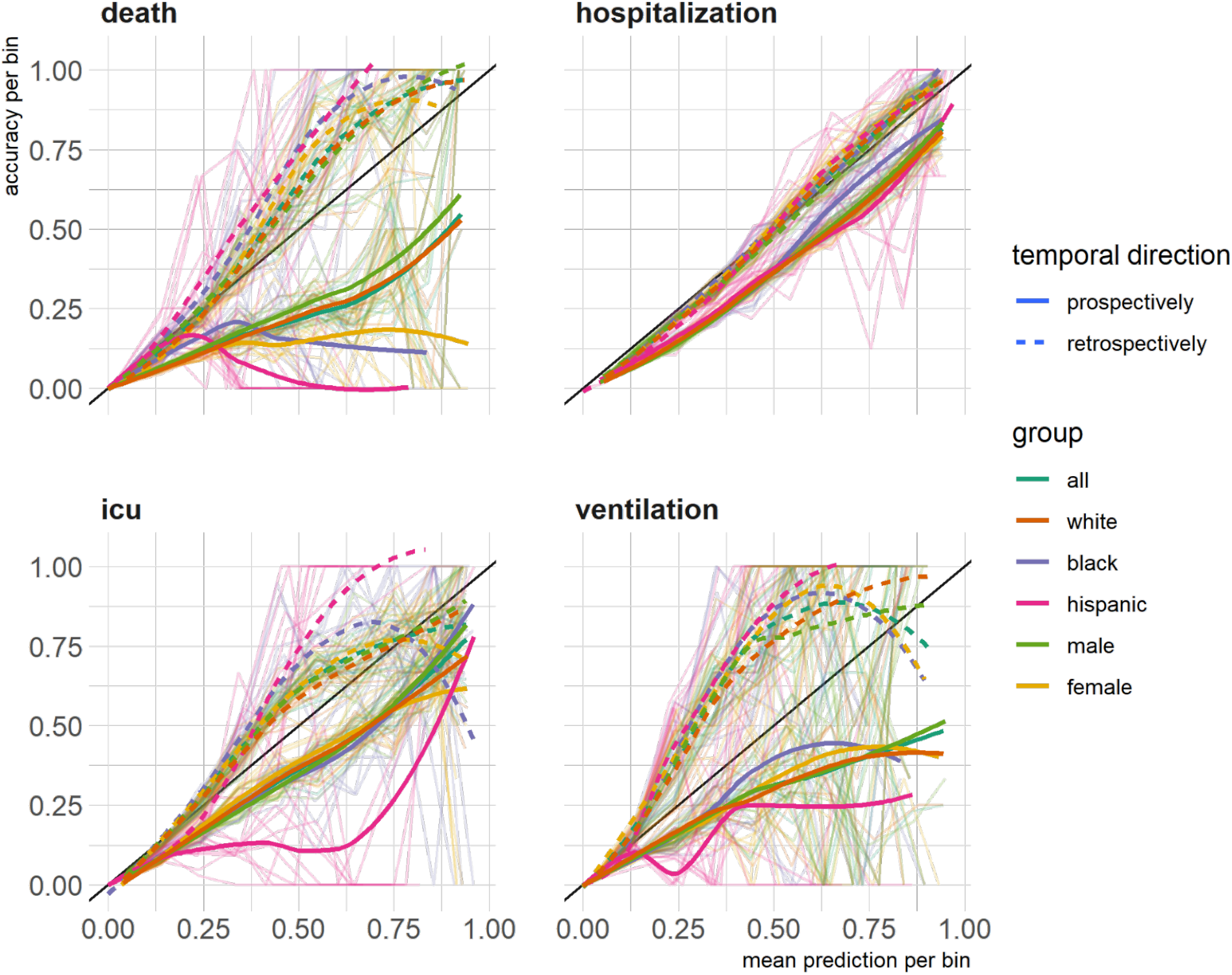
The diagnostic reliability (calibration) diagrams for each outcome broken by group and temporal direction.

Compared to the overall population, retrospectively and prospectively across time, the models marginally performed worse for male patients and better for Hispanic and female patients, as measured by AUROC and higher Brier scores. We use the term “marginal” as the range of delta between performance metrics within demographics groups and the overall model was relatively small. For the rest of the demographic groups, the performances were more mixed. From the diagnostic reliability diagrams, the divergence from the diagonal line is present in three of the four prediction tasks, but there are variabilities across groups in both with no consistent pattern. The only exception in this regard was diminished reliability in prospectively predicted probabilities of COVID-19 mortality among hispanic, female, and black patients.

### Individual-level evaluation

For the individual-level evaluation of the bias, we looked at the mean absolute error across age (Figure 5). We evaluated whether the models’ average error rates (i.e., the absolute difference between the actual outcome and the predicted probabilities) changes as patients’ age increases. An MAE smaller than 0.5 would indicate that the model predicted probability was not far off from the actual outcome. For example, the patient could actually have the outcome and the computed probability would be above 50% and therefore the MAE would be smaller than 0.5. To visualize the trends, we fit a smoothed trendline using generalized additive models. For all outcomes, modeling error seemed to increase as the patients became older, and the patterns were almost identical retrospectively and prospectively. None of the trend lines passed the 0.5 threshold, which means despite the higher error rates for the older patients, the models provide acceptable errors for the majority of the patients. The lowest error rates were observed in predicting ventilation. In the case of predicting mortality and hospitalization, the error rates increasingly escalated by age, whereas in predicting ICU admission and need for mechanical ventilator, error rates peaked at around 75 years and then diminished for older patients.

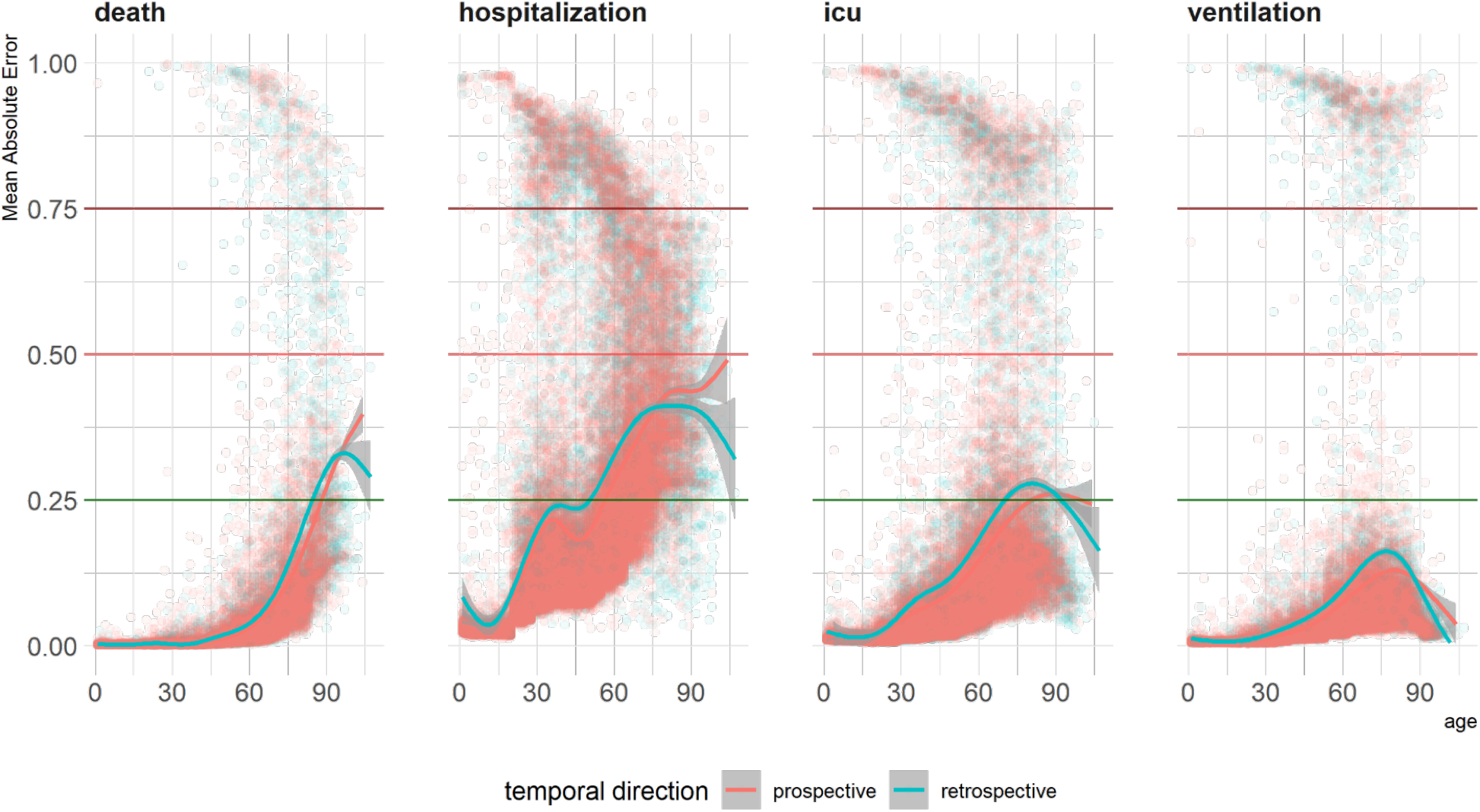

## Discussion

From a model-level perspective, we did not find consistent biased behaviors in predictive models against all minority groups. From the individual-level, we found consistent bias in increasing error rates for older patients. It is known that a predictive model’s reliability (calibration) and discrimination cannot both be maximized simultaneously.^35^ That is, for example, improving reliability may not meaningfully improve discrimination.^39^ Yet, there are ad hoc calibration methodologies to scale predicted probabilities for better clinical interpretation. We argue that proper evaluation of bias in medical AI requires a holistic approach that can invigorate follow-up investigations for identifying the underlying roots of bias, rather than providing a partial perspective that may not lead to constructive improvements.

To an AI algorithm, bias can happen due to the signal strength (or lack thereof) in one or more of the features (i.e., variables, covariates, predictors). That is, the model which has been trained on a certain predictor may not predict well for a certain protected group because the important predictors are not available or are noisy in that population. This, in turn, could have multiple underlying causes, such as healthcare disparities that can influence access to care, systematic inequalities, data quality issues, biological factors, and/or socio-economic and environmental determinants. Some of this bias can be addressed by post-processing techniques, depending on which aspect of bias one aims to address. We concluded that medical AI bias is multi-faceted and requires multiple perspectives to be practically addressed. Nevertheless, the first step for addressing the bias in medical AI is to identify bias in a way that can be traced back to its root.

Compared to the overall population, retrospectively and prospectively across time, the models marginally performed worse for male patients and better for Hispanic and female patients, as measured by AUROC and Brier scores. The range of delta between these performance metrics within demographics groups and the overall model was relatively small. For the rest of the demographic groups, the performances were more mixed. The models’ performance metrics remained stable until June 2021. That is, the models that were developed with data from March to September 2020, provided relatively stable predictive performance prospectively up until May-June 2021. Despite the increased variability, the prospective modeling performance remained high for predicting hospitalization and the need for mechanical ventilators.

COVID-19 vaccinations became widely available in the spring of 2021. It is possible that the widespread use of vaccinations throughout Massachusetts, along with the incorporation of other proven therapies including dexamethasone^40^ and Remdesivir, ^41^ changed outcomes for patients. Also, the case rate in Massachusetts was very low in July,^42^ which may have resulted in increased capacity compared to the outset of the pandemic when the healthcare system was stressed. Additionally, the Delta variant was expected to be the dominant strain of Coronavirus in Massachusetts.^43^ The mutations to the virus itself could potentially change outcomes for patients. While we do not know exactly what led to the decreased performance of the model in July, future studies should consider characterizing whether the model overestimates or underestimates an outcome in certain populations, which could give further insight into how these changes are cumulatively having a favorable or adverse impact on patient care.

From the reliability/calibration perspective, except in the case of prospective evaluation of hospitalization predictions among Hispanic, female, and black patients, the diagnostic reliability diagrams did not show consistent bias towards or against a certain group.

In terms of the mean absolute error between the actual outcome and the estimated probabilities, we did see error rates increase over age, but the error rates were not critical in that one could still assign the patients to the correct group based on the produced probabilities. More continuous metrics need to be evaluated at the patient level for a comprehensive view of changes in AI bias against or towards certain patients.

We evaluated raw predicted scores. There is a large body of work on calibrating prediction scores for improving the reliability of prediction models in clinical settings.^44–47^ Calibration methods are useful ad hoc solutions for increasing the reliability of the prediction models. We show in Figure 3S that isotonic calibration,^48^ for instance, can provide more reliable predictive scores and may reduce bias. However, unless calibration methods are embedded into a predictive modeling pipeline, their impact on improving or aggravating bias in medical AI needs to be fully evaluated as a post-processing step.

Given that we face systemic bias in our country’s core institutions, we need technologies that will reduce these disparities and not exacerbate them.^38^ There are efforts from the larger AI community, such as AI Fairness 360^49^ and Failearn,^50^ to develop open-source software systems for measuring and mitigating bias. These programs are often ad hoc or work as standalone post processing solutions. We plan to compare these model independent methods and add relevant functionalities to our domain specific approach.

The premise for evaluating these predictive models was to create a framework for discovering and quantifying the various types of biases towards different subgroups that were encoded unintentionally. We have incorporated the presented bias measurement framework within the MLHO pipeline,^24^ which is specifically designed for modeling clinical data. We believe that providing means to evaluate and address unrecognized bias within a data-centric pipeline will enable the generation of medical AI that takes into account various biases while in production.

## Data Availability

Protected Health Information restrictions apply to the availability of the clinical data here, which were used under IRB approval for use only in the current study. As a result, this dataset is not publicly available.

## Data Availability Statement

Protected Health Information restrictions apply to the availability of the clinical data here, which were used under IRB approval for use only in the current study. As a result, this dataset is not publicly available. Qualified researchers affiliated with the Mass General Brigham (MGB) may apply for access to these data through the MGB Institutional Review Board.

## Code Availability Statement

The R code to perform this analysis is available at https://hestiri.github.io/mlho

## References

1. Vayena, E., Blasimme, A. & Cohen, I. G. Machine learning in medicine: Addressing ethical challenges. PLoS Med. 15, e1002689 (2018).

2. Char, D. S., Shah, N. H. & Magnus, D. Implementing Machine Learning in Health Care — Addressing Ethical Challenges. New England Journal of Medicine vol. 378 981–983 (2018).

3. Moratinos, G. L., Lazcoz Moratinos, G. & de Miguel Beriain, I. Big Data Analysis and Machine Learning in Intensive Care Medicine: Identifying new ethical and legal challenges. Medicina Intensiva (English Edition) vol. 44 319–320 (2020).

4. Hajjo, R. The Ethical Challenges of Applying Machine Learning and Artificial Intelligence in Cancer Care. 2018 1st International Conference on Cancer Care Informatics (CCI) (2018) doi:10.1109/cancercare.2018.8618186.

5. DeCamp, M. & Lindvall, C. Latent bias and the implementation of artificial intelligence in medicine. J. Am. Med. Inform. Assoc. 27, 2020–2023 (2020).

6. Chouldechova, A. & Roth, A. A snapshot of the frontiers of fairness in machine learning. Commun. ACM 63, 82–89 (2020).

7. Obermeyer, Z., Powers, B., Vogeli, C. & Mullainathan, S. Dissecting racial bias in an algorithm used to manage the health of populations. Science 366, 447–453 (2019).

8. Noor, P. Can we trust AI not to further embed racial bias and prejudice? BMJ 368, (2020).

9. Adamson, A. S. & Smith, A. Machine Learning and Health Care Disparities in Dermatology. JAMA Dermatology vol. 154 1247 (2018).

10. Clarke, Y. D. Algorithmic Accountability Act of 2019. (2019).

11. Floridi, L. et al. AI4People—An Ethical Framework for a Good AI Society: Opportunities, Risks, Principles, and Recommendations. Minds Mach. 28, 689–707 (2018).

12. Klare, B. F., Burge, M. J., Klontz, J. C., Vorder Bruegge, R. W. & Jain, A. K. Face Recognition Performance: Role of Demographic Information. IEEE Trans. Inf. Forensics Secur. 7, 1789–1801 (2012).

13. O’Toole, A. J., Dunlop, J., An, X. & Phillips, P. J. Demographic effects on estimates of automatic face recognition performance. (2011) doi:10.6028/nist.ir.7757.

14. Hupont, I. & Fernandez, C. DemogPairs: Quantifying the Impact of Demographic Imbalance in Deep Face Recognition. 2019 14th IEEE International Conference on Automatic Face & Gesture Recognition (FG 2019) (2019) doi:10.1109/fg.2019.8756625.

15. Caliskan, A., Bryson, J. J. & Narayanan, A. Semantics derived automatically from language corpora contain human-like biases. Science 356, 183–186 (2017).

16. Aran, X. F., Such, J. M. & Criado, N. Attesting Biases and Discrimination using Language Semantics. arXiv [cs.AI] (2019).

17. Rice, J. J. & Norel, R. Faculty Opinions recommendation of Semantics derived automatically from language corpora contain human-like biases. Faculty Opinions – Post-Publication Peer Review of the Biomedical Literature (2017) doi:10.3410/f.727506427.793532942.

18. Rajkomar, A., Hardt, M., Howell, M. D., Corrado, G. & Chin, M. H. Ensuring Fairness in Machine Learning to Advance Health Equity. Ann. Intern. Med. 169, 866–872 (2018).

19. Cormier, J. N. et al. Ethnic differences among patients with cutaneous melanoma. Arch. Intern. Med. 166, 1907–1914 (2006).

20. Kagiyama, N., Shrestha, S., Farjo, P. D. & Sengupta, P. P. Artificial Intelligence: Practical Primer for Clinical Research in Cardiovascular Disease. J. Am. Heart Assoc. 8, e012788 (2019).

21. Lopez-Jimenez, F. et al. Artificial Intelligence in Cardiology: Present and Future. Mayo Clin. Proc. 95, 1015–1039 (2020).

22. Tat, E., Bhatt, D. L. & Rabbat, M. G. Addressing bias: artificial intelligence in cardiovascular medicine. Lancet Digit Health 2, e635–e636 (2020).

23. Parikh, R. B., Teeple, S. & Navathe, A. S. Addressing Bias in Artificial Intelligence in Health Care. JAMA 322, 2377–2378 (2019).

24. Estiri, H., Strasser, Z. H. & Murphy, S. N. Individualized prediction of COVID-19 adverse outcomes with MLHO. Sci. Rep. 11, 5322 (2021).

25. Estiri, H. et al. Predicting COVID-19 mortality with electronic medical records. NPJ Digit Med 4, 15 (2021).

26. Estiri, H., Vasey, S. & Murphy, S. N. Transitive Sequential Pattern Mining for Discrete Clinical Data. in Artificial Intelligence in Medicine 414–424 (Springer International Publishing, 2020).

27. Estiri H, Strasser ZH, Klann JG, McCoy TH Jr., Wagholikar KB, Vasey S, Castro VM, Murphy ME, Murphy SN. Transitive Sequencing Medical Records for Mining Predictive and Interpretable Temporal Representations. Patterns (2020).

28. Mehrabi, N., Morstatter, F., Saxena, N., Lerman, K. & Galstyan, A. A Survey on Bias and Fairness in Machine Learning. ACM Comput. Surv. 54, 1–35 (2021).

29. Chouldechova, A. Fair Prediction with Disparate Impact: A Study of Bias in Recidivism Prediction Instruments. Big Data 5, 153–163 (2017).

30. Verma, S. & Rubin, J. Fairness Definitions Explained. in 2018 IEEE/ACM International Workshop on Software Fairness (FairWare) 1–7 (2018).

31. Brier, G. W. Verification of forecasts expressed in terms of probability. Mon. Weather Rev. 78, 1–3 (1950).

32. Ferro, C. A. T. Comparing Probabilistic Forecasting Systems with the Brier Score. Weather Forecast. 22, 1076–1088 (2007).

33. Benjamini, Y. & Yekutieli, D. The Control of the False Discovery Rate in Multiple Testing under Dependency. Ann. Stat. 29, 1165–1188 (2001).

34. Huang, Y., Li, W., Macheret, F., Gabriel, R. A. & Ohno-Machado, L. A tutorial on calibration measurements and calibration models for clinical prediction models. J. Am. Med. Inform. Assoc. 27, 621–633 (2020).

35. Van Calster, B. et al. Calibration: the Achilles heel of predictive analytics. BMC Med. 17, 230 (2019).

36. Wood, S. Generalized Additive Models: An Introduction with R, Second Edition. (CRC Press, 2017).

37. Wood, S. Package ‘mgcv’. (2021).

38. Kaushal, A., Altman, R. & Langlotz, C. Health Care AI Systems Are Biased. Scientific American (2020).

39. Cook, N. R. Statistical evaluation of prognostic versus diagnostic models: beyond the ROC curve. Clin. Chem. 54, 17–23 (2008).

40. RECOVERY Collaborative Group et al. Dexamethasone in Hospitalized Patients with Covid-19. N. Engl. J. Med. 384, 693–704 (2021).

41. Beigel, J. H. et al. Remdesivir for the Treatment of Covid-19 - Final Report. N. Engl. J. Med. 383, 1813–1826 (2020).

42. Massachusetts Coronavirus Map and Case Count. The New York Times (2020).

43. Markos, M. Delta Variant Taking Over as Dominant Strain in Mass., Experts Say. NBC10 Boston https://www.nbcboston.com/news/coronavirus/delta-variant-taking-over-as-dominant-strain-in-mass-experts-say/2423599/ (2021).

44. Benevenuta, S., Capriotti, E. & Fariselli, P. Calibrating variant-scoring methods for clinical decision making. Bioinformatics (2021) doi:10.1093/bioinformatics/btaa943.

45. Alba, A. C. et al. Discrimination and Calibration of Clinical Prediction Models: Users’ Guides to the Medical Literature. JAMA 318, 1377–1384 (2017).

46. Van Calster, B. & Vickers, A. J. Calibration of risk prediction models: impact on decision-analytic performance. Med. Decis. Making 35, 162–169 (2015).

47. Holmberg, L. & Vickers, A. Evaluation of prediction models for decision-making: beyond calibration and discrimination. PLoS medicine vol. 10 e1001491 (2013).

48. Mair, P., Hornik, K. & de Leeuw, J. Isotone Optimization in R: Pool-Adjacent-Violators Algorithm (PAVA) and Active Set Methods. J. Stat. Softw. 32, 1–24 (2009).

49. Bellamy, R. K. E. et al. AI Fairness 360: An extensible toolkit for detecting and mitigating algorithmic bias. IBM J. Res. Dev. 63, 4:1–4:15 (2019).

50. Bird, S. et al. Fairlearn: A toolkit for assessing and improving fairness in AI. Microsoft, Tech. Rep. MSR-TR-2020-32 (2020).

